# Systematic profiling of SARS-CoV-2 specific IgG epitopes at single amino acid resolution

**DOI:** 10.1101/2020.09.08.20190496

**Authors:** Huan Qi, Ming-liang Ma, He-wei Jiang, Jian-ya Ling, Ling-yun Chen, Hai-nan Zhang, Dan-yun Lai, Yang Li, Zi-wen Guo, Chuan-sheng Hu, Shu-Juan Guo, Qing-feng Meng, Yan Ren, Wei Wang, Xiao Yang, Jie Zhou, Xiao-dong Zhao, Hua Li, Sheng-ce Tao

**Author notes:** These authors contributed equally (Huan Qi, Ming-liang Ma, He-wei Jiang, Jian-ya Ling, Ling-yun Chen). Corresponding Authors, (S.-c. T.), (H. L.), (X.-d. Z.), (J. Z.).

## Abstract

SARS-CoV-2 specific IgG responses play critical roles for patients to recover from COVID-19, in-depth dissecting of the IgG responses on systems level is of great interest. Herein, we adopted a newly developed high-throughput epitope mapping technology (AbMap), analyzed 55 COVID-19 convalescent sera and 226 antibody samples enriched by specific proteins or peptides from these sera. We revealed three areas that are rich of IgG epitopes, two are on Spike protein but outside of RBD, and one is on Nucleocapsid protein. We identified 29 significant epitopes on Spike protein, from two of these significant epitopes, two critical epitope residues were found, *i. e*., D936 and P1263, which are highly related to the infectivity of SARS-CoV-2 In summary, we provided the first global map of IgG binding epitopes for SARS-CoV-2 at single amino acid resolution. This map will facilitate the precise development of therapeutic antibodies and vaccines.

**HIGHLIGHTS:**

1. A map of SARS-CoV-2 specific IgG binding epitopes at single amino acid resolution
2. Two areas outside of RBD that are rich of significant epitopes were identified
3. One area rich of significant epitopes was determined on Nucleocapsid protein
4. Two critical epitope residues (D936 and P1263) on Spike protein are highly related to the infectivity of SARS-CoV-2

## INTRODUCTION

COVID-19 is a worldwide pandemic and caused by SARS-CoV-2 (*1, 2*). By September 8, 2020, 27,249,308 cases were diagnosed and 890,971 lives were claimed (https://coronavirus.jhu.edu/map.html) (*3*). Vaccines and therapeutic antibodies of high effectivity are desperately needed. It is known that SARS-CoV-2 encodes 27 proteins, of these proteins, Spike protein (S protein) plays a critical role in the specific binding of SARS-CoV-2 to human cells. During the infection, specific proteases cut S protein at S2’ and furin site to split S1 and S2 (*4*). S protein is rich of N-glycosylation, *i. e*., 21 sites (*5*). On S protein, RBD domain is presently the central target to develop SARS-CoV-2 antibodies and vaccines (*6–10*). Other areas of S protein can also elicit B cell responses and neutralization antibodies (*11–15*). Nucleocapsid protein (N protein) is highly abundant and triggers significant IgG response (*16–18*).

SARS-CoV-2 specific IgG response is critical for the recovery of COVID-19 patient, since specific and effective therapy is still not available. The comprehensive and in-depth revealing of SARS-CoV-2 specific IgG responses will definitely help us to understand the COVID-19 immunity better, and to facilitate the precise development of neutralization antibodies and vaccines. To reveal the SARS-CoV-2 specific IgG response, several studies have been performed through different angles by applying different strategies. By analyzing 55 convalescent sera on a SARS-CoV-2 proteome microarray, the IgG responses against 18 SARS-CoV-2 proteins were surveyed, including S1, S2, N and RBD (*16*). By analyzing sera from 1,051 patients on a S protein peptide microarray of full S protein coverage, several areas and a set of peptides of high IgG reactivity were identified (*14*). Similar studies were also performed by using another peptide microarray (*17*), and a coronavirus specific phage displayed peptide library (*19*). The IgG responses on protein and peptide levels, but not single amino acid level, were at least partially revealed by these studies.

Herein, we aimed to go one step further to dissect the SARS-CoV-2 specific IgG responses systematically at single amino acid level. We adopted AbMap, a method that was developed recently (*20*) for high-throughput epitope mapping. A set of 55 convalescent sera and 226 samples of protein/ peptide enriched antibodies from these sera were analyzed. A map of SARS-CoV-2 specific IgG epitopes at single amino acid resolution was generated for the first time.

## RESULTS

### Schematic diagram: an epitope mapping strategy with three layers of samples

To globally profile the IgG binding epitopes on SARS-CoV-2 proteins from COVID-19 sera, we collected sera from 55 convalescent patients, and 25 controls. The patients were hospitalized in Foshan Fourth hospital in China from 2020-1-25 to 2020-3-8 for various durations. Patient information is summarized in **Table S1**. Serum from each patient was collected on the day of hospital discharge when standard criteria were met.

It is known that serum is rich of antibodies (*7, 21*), and it is reasonable to argue that SARS-CoV-2 specific IgGs will compose only a small portion of the total IgGs in serum. In order to increase the signal to noise ratio of the following-up epitope mapping, the sera from the 55 convalescent patients were subjected for antibody enrichment by specific SARS-CoV-2 proteins and peptides. The three layers of samples, *i. e*., the serum, the protein enriched antibodies, and the peptide enriched antibodies were then subjected for IgG binding epitope mapping by AbMap, a technology that we have established recently (*20*). Statistically significant epitopes were successfully identified for all these three layers of samples (**Fig. 1**). A variety of epitopes identified from the protein and peptide enriched antibodies could be matched to the sequence of the corresponding proteins and peptides. These results indicate AbMap is suitable for dissecting the SARS-CoV-2 IgG binding epitopes using COVID-19 sera as input.

**Figure 1.**
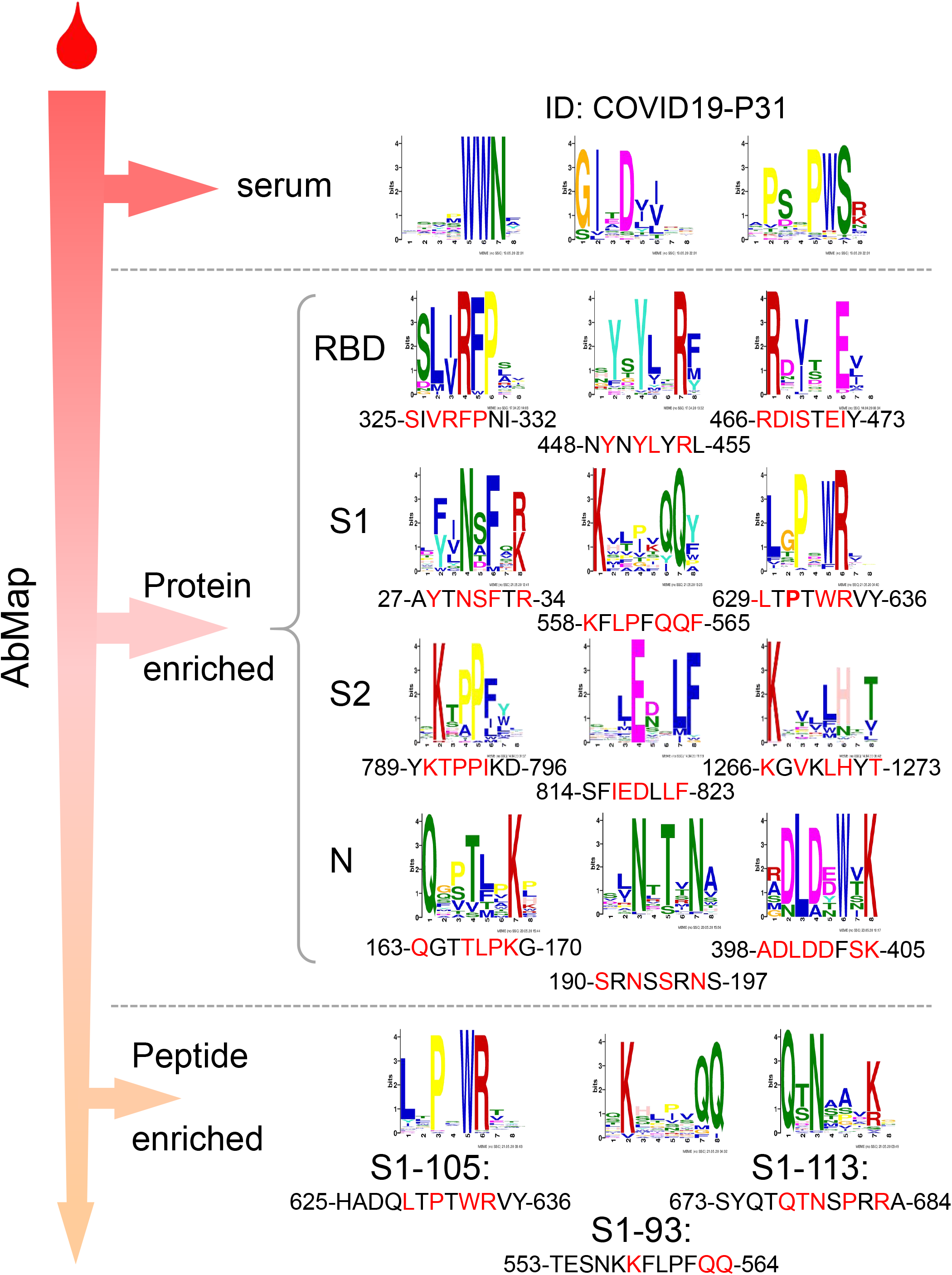
The schematic diagram of using AbMap to determine SARS-CoV-2 IgG binding epitopes. Three types of samples, *i.e*. sera, protein specific antibodies enriched from sera, and peptide/ epitope specific antibodies enriched from sera, were subjected for epitope mapping using AbMap technology. Significant epitopes were enriched for all these three layers of samples. The matched sequences on the targeting proteins were shown for protein and peptide enriched antibodies.

### A map of SARS-CoV-2 specific IgG binding epitopes

To reveal the epitopes at systems level, the 55 convalescent sera were subjected for AbMap analysis directly, following the procedure that we established recently (*20*) and used a high stringent cutoff, we have identified 418 motifs (**Table S2**), 275 of which could be matched to 27 of the 28 known SARS-CoV-2 proteins (**Table S3, Fig. 2A**), we recognized these motifs as epitopes of high confidence. Vertically (**Fig. 2A**), to estimate the variation of SARS-CoV-2 specific IgG responses among the 55 convalescent sera, we added up the epitope frequency for each patient. The results clearly showed that there are significant differences among the patients, with a frequency from 0 to 27. According to the histogram of the frequency (**Fig. 2B**), we noticed that the number of patients is inversely proportional to the frequency, this suggest the SARS-CoV-2 specific IgG response is not randomly distributed among the patients, the underlying mechanism may be worth further exploration. Horizontally (**Fig. 2A**), to check the distribution of the epitopes among the proteins, we also counted the epitopes for each protein, *i. e*., sum_epitopes. There are big variations of the sum_epitopes from 0 for ORF6 to 125 for NSP3 (**Fig. 2C**). Since the size of the 28 SARS-CoV-2 proteins varied a lot from 22 aa (ORF3b) to 1,945 aa (NSP3), it is possible that the variations of the sum_epitopes may be due to the different sizes of the proteins. To check this possibility, we divided sum_epitopes by the length of the proteins, indeed, the variation of the sum_epitopes among protein was largely normalized. However, there are still significant variations for sum_epitopes/ length among the proteins, from 0 for ORF6 to 0.08 for NSP9, these differences may be due to the accessibility of these proteins to the immune system during infection, further study may be needed to address this.

**Figure 2.**
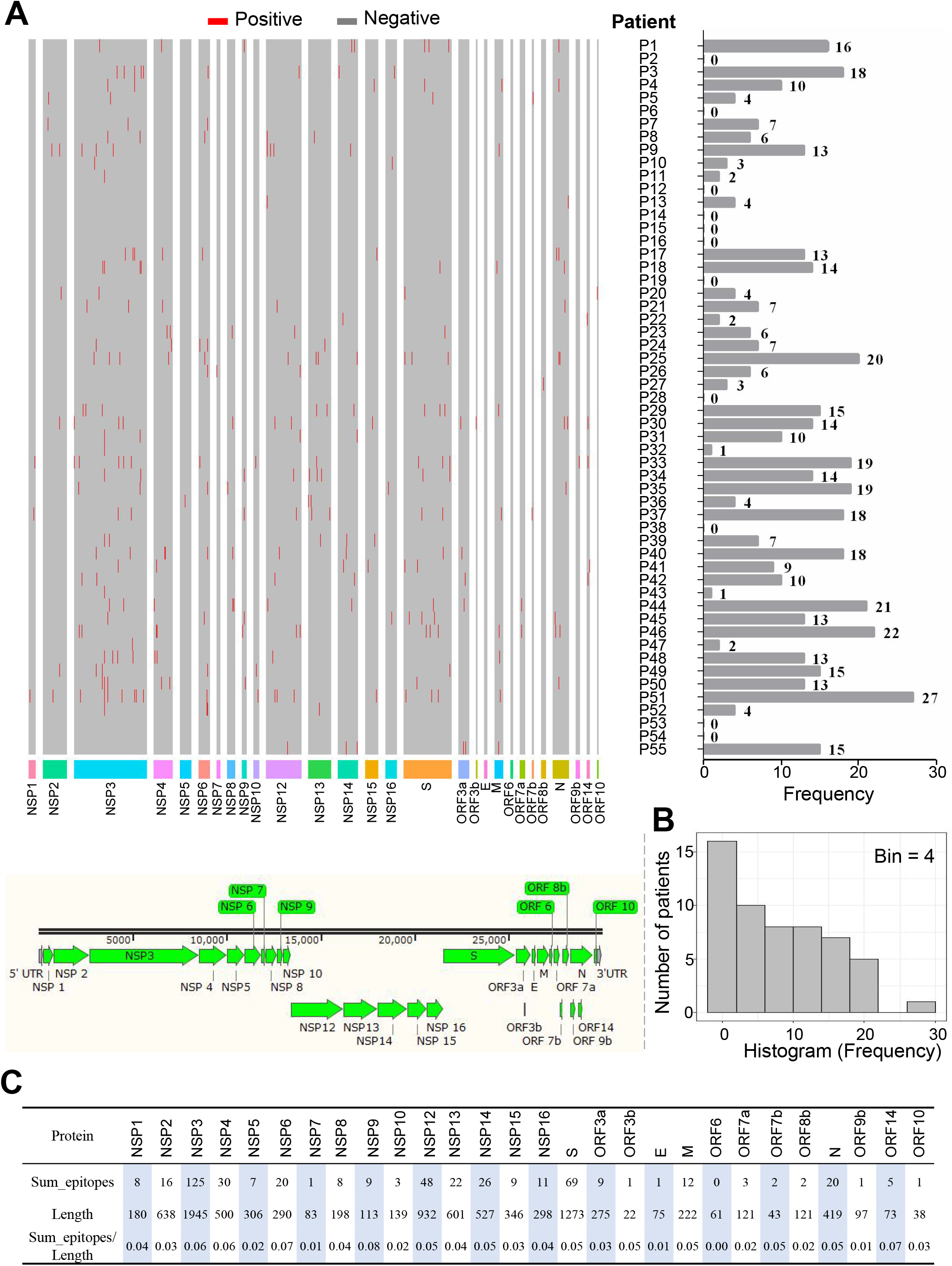
The map of SARS-CoV-2 specific IgG binding epitopes revealed from convalescent sera directly. (A) The epitopes (**Table S3**) of each patient (P1-P55) were plotted alongside the known SARS-CoV-2 proteins from N terminal to C terminal. The total number of the identified epitopes for each patient and each protein were defined as frequency and sum_epitopes, respectively. (B) The histogram of the epitope frequency (bin = 4). (C) The sum_epitopes of each protein.

Except the SARS-CoV-2 specific antibodies, there are many other antibodies in the convalescent sera, it is possible that a big portion of the 143 un-matched motifs may correspond to the un-related antibodies (*22*). To reveal more SARS-CoV-2 epitopes of high confidence, it is necessary to improve the signal-to-noise ratio of AbMap. For this purpose, we decided to enrich protein specific antibodies for AbMap. According to one of our previous study (*16*), we knew N protein, S1, S2 and RBD are of high reactivity among patients. Because we only had very limited amount of sera, we decided to enrich the antibodies from each sample in a consequential manner, *i. e*., RBD, S1, S2 and N. To ensure the quality of the enrichment, proteins that biotinylated through Avi-tag and streptavidin coated magnetic beads were adopted (**Fig. S1**). Clearly, a variety of highly confident epitopes were identified and could be matched to the sequences of the corresponding proteins (**Table S3**).

Thus, we generated a map of SARS-CoV-2 specific IgG binding epitopes by analyzing 55 convalescent sera and 226 samples of antibodies enriched by specific proteins from these sera.

### Two areas outside of RBD that rich of significant epitopes were identified on Spike protein

S protein is the key protein for infection and is known to be highly immunogenic (*15, 16*). To present the distribution of the epitopes on S protein, we merged the sequence matched epitopes revealed from both S1 and S2 enriched antibodies (**Table S3**). When matching the epitopes to S protein sequence, if an epitope sequence obtained from one sample is the same for other samples, we counted this epitope as 1 for this sample and added it up for all the samples as the final frequency of this epitope. We then plotted the epitopes, the epitopes sequences, and the frequencies alongside the linear sequence and domains of S protein (**Fig. 3A**). The results clearly showed that the epitopes are distributed cross but not evenly on S protein. Two hot areas that rich of significant epitopes were readily identified, the first area almost covers the entire CTD (C terminal domain), the second area covers the S2’ protease cleavage site and the fusion peptide (FP). Significant epitopes were also identified on the cytoplasmic C-terminal end of S protein. These results are highly consistent to a peptide microarray-based study, of which about 4,000 samples were analyzed against a set of 197 peptides that covers the entire S protein (*14*, *15*).

**Figure 3.**
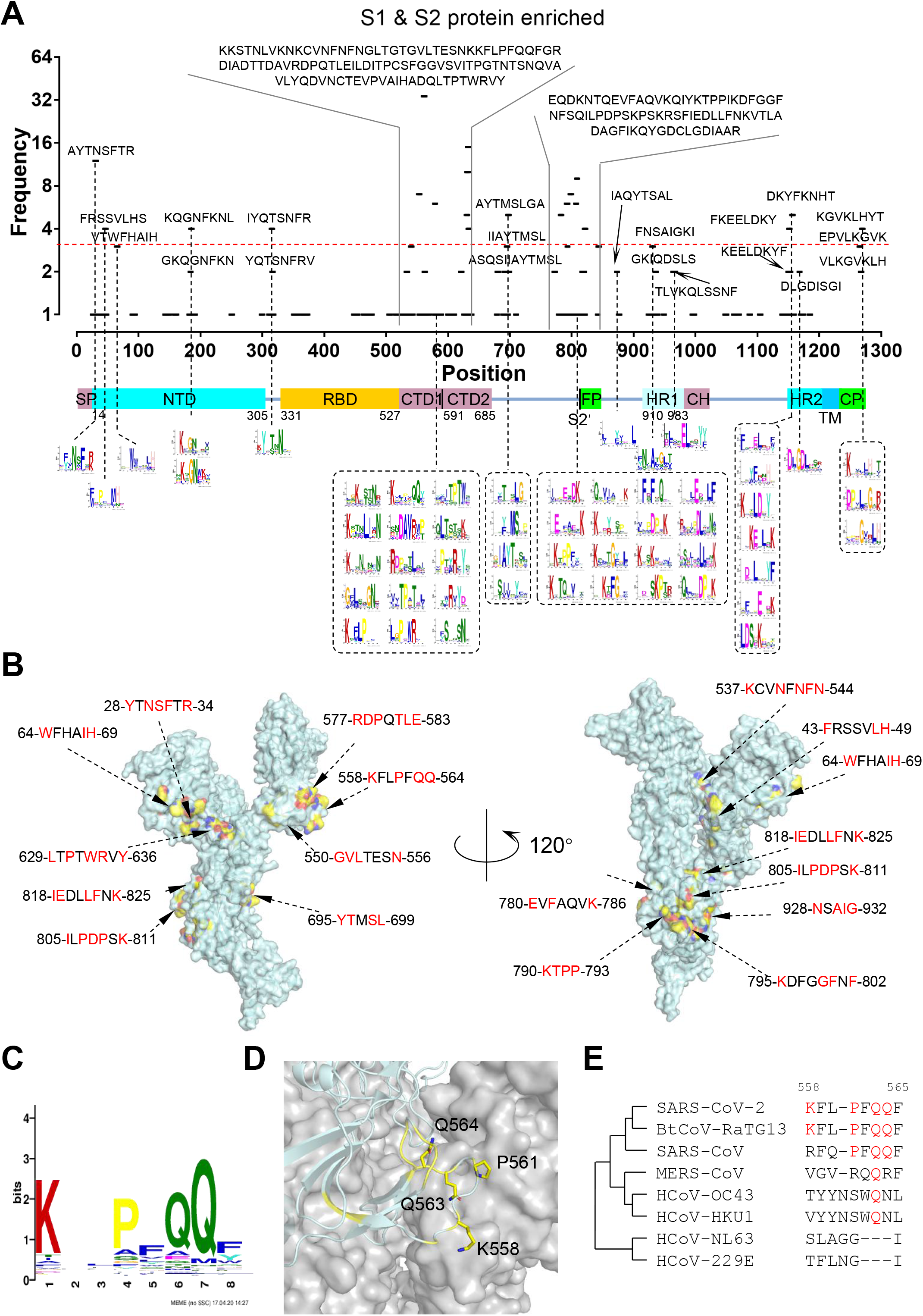
Significant epitopes on S protein. (A) The distribution of the sequence-matched epitopes on S protein. (B) The distribution of the significant epitopes (frequency> = 3) on the monomer 3D structure of the S protein. Red marked amino acids represent key residues of epitopes. (C) A representative significant epitope. (D) The epitope was matched to S protein, critical residues were labeled as yellow. (E) Homology analysis of the epitope among three deadly coronaviruses, *i. e*., SARS-CoV-2, SARS-CoV and MERS-CoV, four common human coronaviruses, *i. e*., HCoV-OC43, HKU-1, NL63 and 229E and the highly homologous bat coronavirus BtCoV-RaTG13.

It is interesting to test the necessity of enriching SARS-CoV-2 specific antibodies from sera. To this end, we took S protein as an example, we plotted the epitopes identified by using serum directly (**Fig. S2** black dots) and the S protein enriched antibodies (**Fig. S2** red dots) alongside the sequence of S protein. The distributions are similar between sera and S protein enriched antibodies, as expected, many more significant epitopes with higher frequency were identified by using the S protein enriched antibodies (**Fig. S2**). This improvement may be due to the removal of unrelated antibodies from sera, thus the signal-to-noise ratio was significantly enhanced.

We defined the epitopes with frequency > = 3 as significant epitopes and obtained 28 of them (**Fig. S3A**). To further illustrate the location and distribution of the 28 significant epitopes (**Fig. S3A**), we mapped them to the 3D structure of Spike protein (*23*) monomer (**Fig. 3B**) and trimer (**Fig. S3B**). It is clear that most of these epitopes locate on the surface of Spike protein, which is consistent to the common notion (*24*). However, Epitope-S1-6 (KCVNFNFN) and Epitope-S2-3 (QEVFAQVK) are not on the surface of the trimeric Spike protein, but on the surface of the monomer. A plausible explanation is that specific areas of the Spike protein monomer could be exposed to the immune system during the infection. To further pinpoint the locations of the critical residues of the significant epitopes, we took 4 critical epitope residues, *i. e*., K558, P561, Q563 and Q564 from a significant epitope as examples (**Fig. 3C**). As expected, all these residues locate on the surface of S protein (**Fig. 3D**). To check the similarity of the epitopes/ critical epitope residues among human coronaviruses, we performed homology analysis for Epitope-S1-8 (KFLPFQQF) (**Fig. S3A**). High homologies were observed among SARS-Cov-2, SARS-CoV and BtCoV-RaTG13 (**Fig. 3E**). Structure analysis and homology analysis were also performed for all the rest 27 significant epitopes wherever applicable (**Table S4**). Similar to that of Epitope-S1-8 (KFLPFQQF), most of the critical epitopes of these 27 significant epitopes also locate on the surface of S protein. High homologies were also observed among SARS-Cov-2, SARS-CoV and BtCoV-RaTG13 for most of the epitopes. In addition, an epitope of high homology among BtCoV-RaTG13, the three deadly human coronaviruses (SARS-CoV-2, SARS-CoV and MERS-CoV) and four common human coronaviruses (HCoV-OC43, HKU1, NL63 and 229E) was also identified, *i. e*., FxxELxxY (**Table S4**).

These results indicate that the critical epitope residues of the significant epitopes are unevenly distributed on S protein both linearly and conformationally. And the critical epitope residues are highly homologous among the 7 human coronaviruses, especially between SARS-CoV-2 and SARS-CoV.

### Critical epitope residues related to the binding of Spike protein and ACE2/ neutralization antibodies, and the infectivity of SARS-CoV-2 were revealed

Because the central role that RBD plays in binding of SARS-CoV-2 to the host cell, biotinylated RBD was also applied to enrich specific antibodies from COVID-19 convalescent sera (**Fig. S1**). One relative significant epitope was identified (**Fig. 4A**). This epitope is also consistent to our peptide microarray-based study (*14*). To visualize the conformational location of this epitope, we mapped the critical epitope residues (yellow), *i. e*., R466, D467, I468, S469, E471 and E473 to the 3D structure of RBD (*25*) (**Fig. 4B**). It is clear that the critical epitope residues are not on the exact binding interface of RBD and ACE2 but adjacent (**Fig. 4C**). To further investigate whether these critical residues interference with the binding of neutralization antibodies to RBD, we studied the co-crystal structures of RBD bound by several well-studied neutralization antibodies, *i. e*., CR3022-Fab (*8*) (**Fig. 4D**), S309-Fv (*26*) (**Fig. 4E**), P2B-2F6-Fab (*27*) (**Fig. S4A**), BD23-Fab (*7*) (**Fig. S4B**), and CB6-Fab (*28*) (**Fig. S4C**). Interestingly, the critical residues are also not on the exact binding interface of RBD and these neutralization antibodies but adjacent. The antibodies targeting this epitope may cause conformational change or providing spatial hinderance, and thus interference the binding of ACE2 and these neutralization antibodies to RBD.

**Figure 4.**
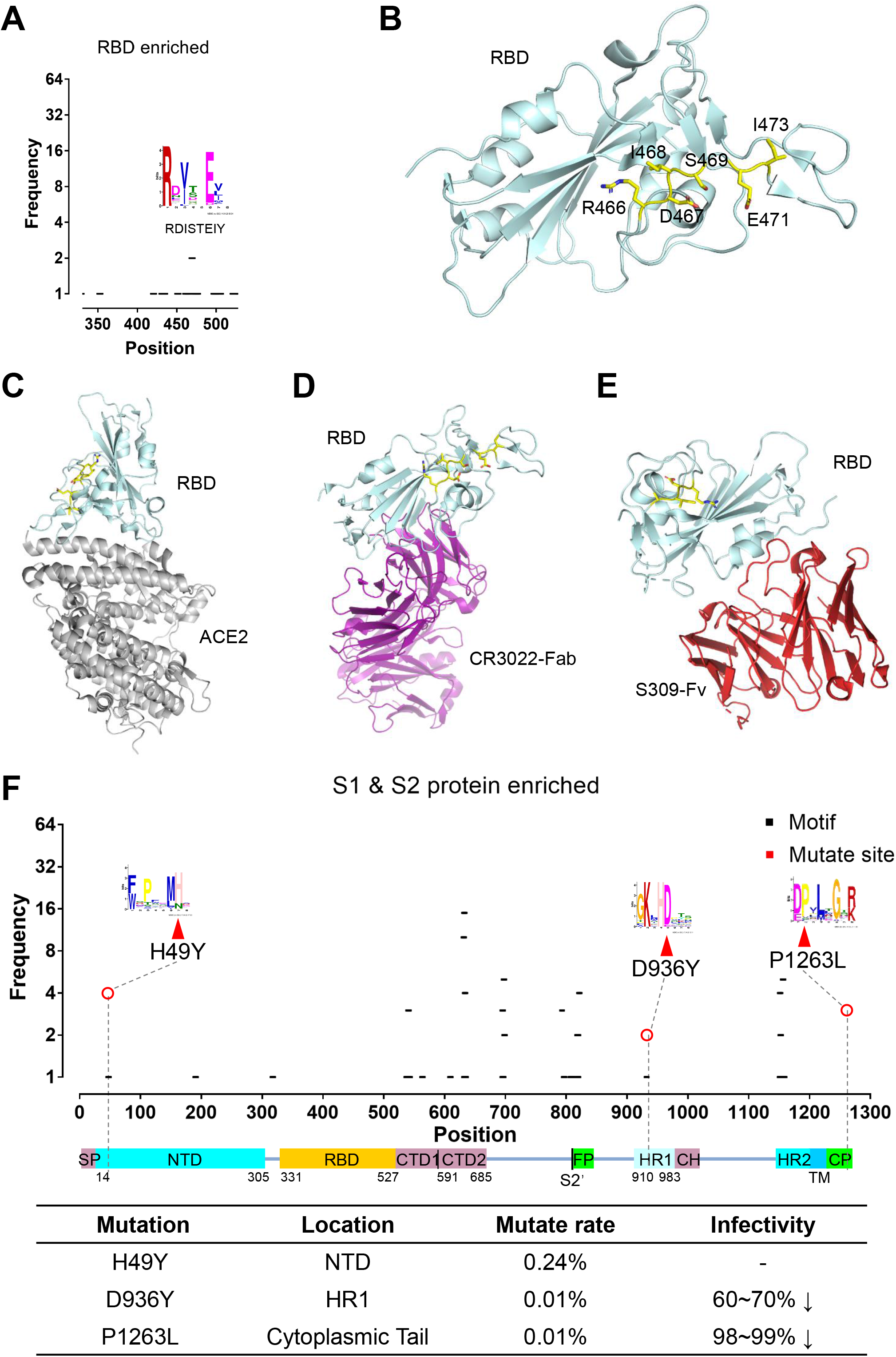
Functional analysis of several key sites of the significant epitopes of S protein. (A) One relatively significant epitope (frequency > = 2) was identified for RBD enriched antibody. (B) Match the critical epitope residues to the structure of RBD. (C) The critical epitope residues locate adjacent to but not at the binding interfaces of RBD and ACE2. (D and E) The critical epitope residues locate adjacent to but not at the binding interfaces of RBD by two highly potent neutralization antibodies, CR3022-Fab(D) and S309-Fv (E). (F) Match the epitopes on S protein with the critical naturally existing mutants (*29*).

Some of the epitopes and the critical epitope residues may be related to the proper function of S protein and the infectivity of SARS-CoV-2. A recent study surveyed the functional roles of a variety of SARS-CoV-2 S protein mutants, and identified a set of point mutations that cause significant reduction of the infectivity (*29*). To explore the roles of the critical epitope residues, we matched the mutation sites with the epitope map of S protein (**Fig. 3A**), we found three mutation sites are also critical epitope residues, *i*. *e*., H49Y, D936Y and P1263L. Except H49Y, D936Y and P1263L could cause 60-70% and 98-99% infectivity loss, respectively. This strongly suggest that D936 and P1263 are critical for maintaining the proper activity of S protein. According to the 2019 Novel Coronavirus Resource (2019nCoVR) database of China National Center for Bioinformation (CNCB) (https://bigd.big.ac.cn/ncov/), those three mutation sites are also of high frequency (**Fig. S4D**). Taken together, it is reasonable to argue that D936 and P1263 may could serve as very promising targets for neutralization antibody and vaccine development.

### One area rich of significant epitopes was determined on Nucleocapsid protein

N protein is highly abundant (*30, 31*) and triggers profound IgG response in COVID-19 patients (*18, 32*). To present the distribution of the epitopes on N protein, we matched the epitopes to N protein sequence. Following the same procedure as that of S protein (**Fig. 3A**), we calculated the frequency of each epitope, and plotted the epitopes, the epitopes sequences, and the frequencies alongside the linear sequence and domains of N protein. The results clearly showed that the epitopes are distributed cross but not evenly on N protein. One hot area rich of significant epitopes were readily identified (**Fig. 5A**), and 6 significant epitopes were identified when set the cutoff as frequency > = 3 (**Fig. 5B**). Because the intact 3D structure of N protein is still not available, we thus decided only to perform homology analysis for N protein. High homologies for the epitopes and the critical epitope residues were observed among SARS-Cov-2, SARS-CoV and BtCoV-RaTG13 for all the 6 significant epitopes (**Fig. 5C**).

**Figure 5.**
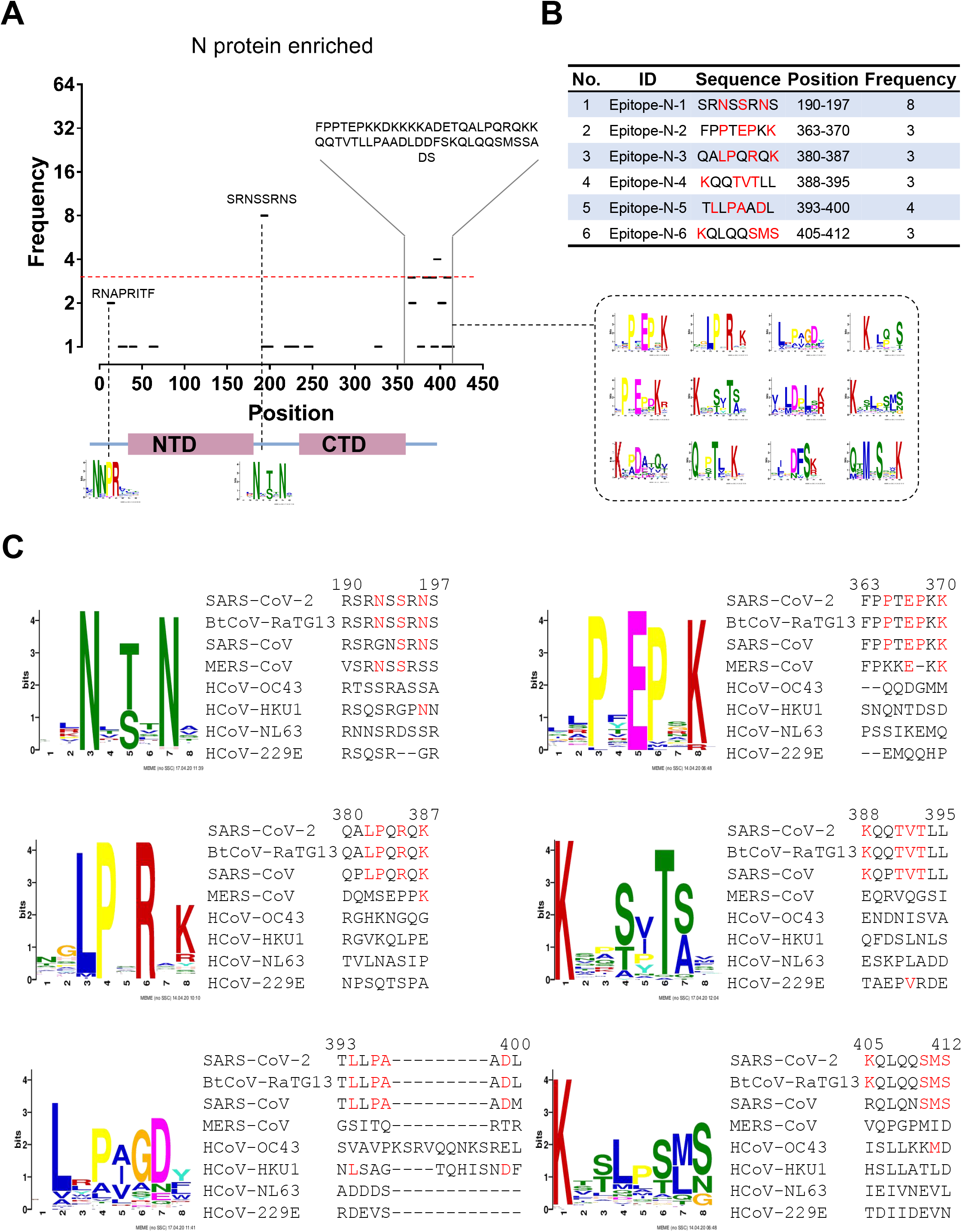
Significant epitopes on N protein. (A) The distribution of the sequence-matched epitopes on N protein. (B) The list of significant epitopes (frequency > = 3). (C) Homology analysis of the significant epitopes among three deadly coronaviruses, *i. e*., SARS-CoV-2, SARS-CoV and MERS-CoV, four common human coronaviruses, *i. e*., HCoV-OC43, HKU-1, NL63 and 229E and the highly homologous bat coronavirus BtCoV-RaTG13.

### Significant epitopes were validated by alanine scanning using a peptide microarray

To validate the key residues of the significant epitopes, according to the frequency and the critical role of RBD, we selected four epitopes, *i. e*., Epitope-S1-8 (KFLPFQQF) and Epitope-S1-12 (LTPTWRVY), which are of the highest frequency, Epitope-RBD-1 (RDISTEIY) from RBD, and an epitope of low frequency, *i. e*., Epitope-S2-15 (KGVKLHYT) was included as control. We then mutated the amino acids of these epitopes one by one to alanine, synthesized the mutated peptides, conjugated to BSA, and fabricated a peptide microarray (**Fig. S6**). To validate the epitopes on the peptide microarray, we selected 5 sera, from which at least one of the significant epitopes among Epitope-S1-8 (KFLPFQQF), Epitope-S1-12 (LTPTWRVY) and Epitope-RBD-1 (RDISTEIY) was identified by AbMap (**Fig. 6A**). The selected sera were then probed on the peptide microarray. For all the three epitopes, when the corresponding samples were tested, significant binding signal loss was observed when any of the critical residue was mutated to alanine. For example, for P44 and Epitope-S1-8 (KFLPFQQF), when any of the four critical residues was mutated, the signal was almost completely lost (**Fig. 6B**). As expected, this is not the case for the unrelated samples (**Fig. 6B**, black caption). However, the binding intensities varied significantly among samples and epitopes.

**Figure 6.**
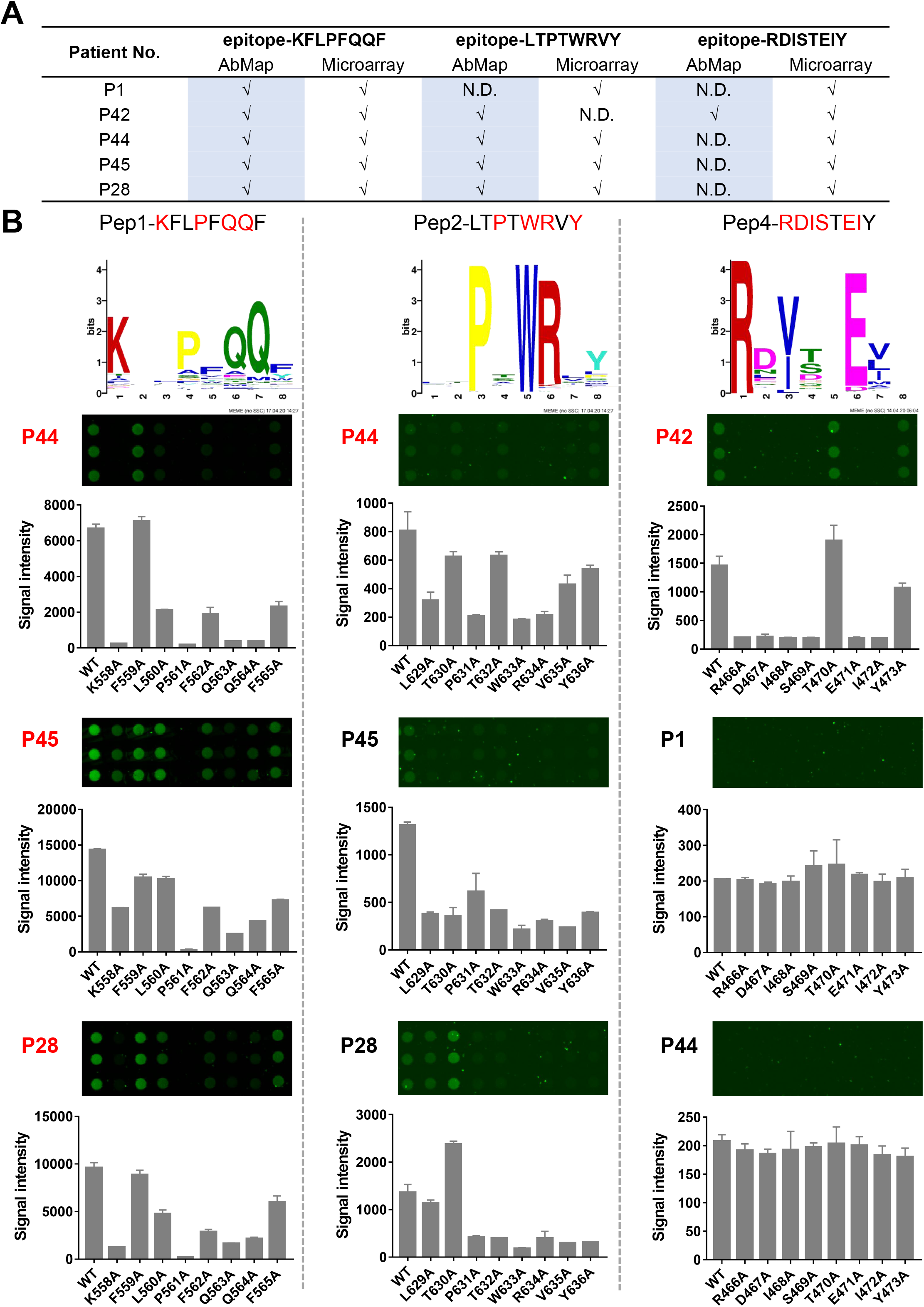
Validation of epitopes by a peptide microarray. (A) Selected samples were subjected for peptide microarray validation. The summary of the validation results. (B) The microarray results of selected samples against three representative epitopes. Sample with red label indicates the corresponding epitope was identified from this sample by AbMap. While other samples were included as negative controls for corresponding epitopes.

It is interesting to see whether the enrichment of SARS-CoV-2 specific antibodies by proteins will enhance the specificity and signal intensity, when probed on the peptide microarray. To test this, we chose Epitope-S1-8 (KFLPFQQF) as an example, probed sera and the corresponding S1 enriched antibodies side by side. As demonstrated in **Fig. S5**, specific bindings were obtained for all the samples. And as expected, the antibody enrichment procedure could significantly enhance both the specificity and the signal intensity. These results further confirmed the necessity for the enrichment of antibodies from sera by specific proteins for epitope mapping.

Taken together, by combining alanine scanning and peptide microarray, the significant epitopes and critical epitope residues could be effectively validated in a fast way.

## DISCUSSION

The aim of this study is to promote our understanding of SARS-CoV-2 specific IgG response in a more precise way. We adopted AbMap technology (*20*), analyzed 55 sera and 226 antibody samples enriched from these sera by RBD, S1, S2 and N proteins. Based on these results, we constructed the first map of SARS-CoV-2 specific IgG binding epitopes at single amino acid resolution.

In this study, by applying AbMap, we developed a two-step strategy for profiling pathogen specific epitopes at single amino acid resolution from sera. IgG response is a central part of B cell immunity. Serum or plasma are rich of IgGs for numerous targets that the immune system has encountered, especially when our body is infected by pathogens (*22*). There is great interest to profile pathogen specific IgG responses by using serum or plasma because of the convenience of sample collection. Protein microarray could be applied to reveal the pathogen specific IgG responses at protein level (*16, 33*), while peptide microarray (*15*) or pathogen specific peptide library (*19, 22*) were able to provide information at peptide/epitope level. However, these platforms could not provide epitope information at amino acid level, and pathogen specific protein or peptide library need to be constructed individually, which is costly and time-consuming. Alternatively, random peptide libraries were attempted for dissecting pathogen specific IgG responses from sera (*34, 35*). For SARS-CoV-2, following the protocol of testing serum directly that we established (*34*), and the K-TOPE algorithm (*35*), we tried to profile the virus specific IgG responses at proteome level. However, most of the identified epitopes that match the sequence of SARS-CoV-2 proteins are of low frequency. This indicates a more powerful algorithm is needed to dig meaningful epitopes for the random peptide-based strategy. We then simplified the data analysis part by combining MEME and FIMO (*36*, *37*), and identified a set of epitopes of marginly statistical significance (**Fig. 1**). To experimentally improve the performance of AbMap, we tried to reduce the noise by enriching SARS-CoV-2 specific IgGs using specific proteins or peptides, a larger number of high confident epitopes were identified as compared to that of testing serum directly. Our results clearly point out the directions to improve phage displayed random peptide library-based strategies, such as AbMap, for pathogen specific epitope mapping: 1. Enrich IgGs by protein/ peptide of interest, and 2. Develop algorithm of better performance.

The epitopes that we identified in this study are highly consistent with studies from our group and others. As expected, S protein and N protein are among the top list according to IgG responses (*16*). High reactivities were also observed for other proteins, for example, NSP3 and NSP12. The different reactivities among the SARS-CoV-2 proteins are correlated to the protein size, but only partially. For example, after normalized by protein size, the highest reactivity was observed for NSP9 (**Fig. 2C**). It is expected that there are other parameter that cause the difference of IgG reactivity, such as protein level (*30, 31*), further study is needed to investigate this.

At peptide/epitope level, our results are also consistent to other studies. Though using a totally different strategy, we found RBD is lack of, while CTD is rich of linear epitope, the other “hot area” of linear epitope is around the S2’ protease cutting site and the fusion peptide. These results are highly consistent to that of the S protein peptide microarray that we constructed (*15*). According to a SARS-CoV-2 specific peptide library based study (*19*), the highly immunogenic regions of Spike protein were also enriched in CTD, region around S2’ and the fusion peptide, and HR2, which are highly consistent to our study. While aa362-399 of N protein which identified as high immunogenic region was also determined in our study, but other highly immunogenic regions were not. The inconsistence may be due to the different sample size and the differences of the detailed protocol. According to a peptide microarray (*17*), 3 of the 6 Spike protein epitopes identified by microarray for serum IgG were also determined as significant epitopes in our study, and aa366-400 of N protein which identified as a long epitope for serum IgG was also enriched in our study. Compared to these epitope related studies, our study can serve as an independent cross validation because of the high consistence. Furthermore, this study is the first of such that providing global IgG epitope information at single amino acid level.

There are a variety of possible applications for the significant epitopes/ critical residues identified in this study. According to S protein, most of the significant epitopes (**Fig. S3A**) are on the surface of the 3D structure, it is reasonable to argue that the significant epitopes (including that from S protein) are naturally of high immunogenicity. Thus, for a given application, one can select one or a few epitopes to generate site specific antibody on demand for S protein as well as other proteins. High frequencies were observed for some of the epitopes, *e. g*., Epitope-S1-8 (KFKPFQQF), it is possible this epitope could be used alone or in combination with other epitope for diagnostics. It is known that areas other than RBD on S protein (*13, 38*), and furthermore, other than RBM on RBD (receptor binding motif) (*8, 39*) may also elicit potent neutralization antibodies. Once determined, the corresponding epitopes could be used directly as vaccine or as immunogen for generating site-specific neutralization antibody. Since the COVID-19 pandemic is unfolding and the sequenced genomes of SARS-CoV-2 is piling up (https://bigd.big.ac.cn/ncov/), undoubtedly, more functionally important amino acid sites will be identified, we can match those sites to the identified critical epitope residues, thus link mutation or SARS-CoV-2 evolution with immune responses. Except for the applications that we discuss here, it is anticipated that more applications of the identified significant epitopes/ critical epitope residues could be explored.

Taken together, we constructed the first map of SARS-CoV-2 specific IgG binding epitopes at single amino acid resolution. We identified several critical epitope residues on S protein that are highly correlate to the infectivity of SARS-CoV-2, and the binding of ACE2/ neutralization antibodies to Spike protein. These results facilitate the in-depth understanding of SARS-CoV-2 specific IgG responses, provide direct hints for precise development of diagnostic reagents, therapeutic antibodies and even vaccines.

## Data Availability

All data supporting the findings of this study could be found within the paper and its Supplementary information, and are available from the related databases. The raw data of NGS were deposited in the NCBI SRA Archive (https://www.ncbi.nlm.nih.gov/sra) with BioProject accession number: PRJNA660911. The raw data of peptide microarray were deposited in the Protein Microarray Database (PMD) (http://www.proteinmicroarray.cn/) with experiment accession number: PMDE246.

## ACKNOWLEDGEMENTS

This work was partially supported by National Key Research and Development Program of China Grant (No. 2016YFA0500600), National Natural Science Foundation of China (No. 31970130, 31670831, 31370813 and 31501054), Open Foundation of Key Laboratory of Systems Biomedicine (No. KLSB2020QN-05).

## AUTHOR CONTRIBUTIONS

S.-c. T., X.-d. Z., and H. L. conceived the idea and designed the study; H. Q. performed the epitope mapping of sera and the protein enriched antibodies; M.-l. M. performed the experiments related peptide microarray; H.-w. J., L.-y. C., and H.-n. Z. isolated the SARS-CoV-2 specific antibodies from sera; Z.-w. G., C.-s. H., X.-d. Z., and H. L. performed NGS and the data analysis; J.-y. L., L.-y. C, S.-q. L, X. Y, W. W., J. Z. provided key reagents. All the authors analyzed the data; S.-c. T, H. Q., and M.-l. M. wrote the manuscript. All the authors have read and approved the final manuscript.

## DECLARATION OF INTEREST

The authors declare no conflict of interest.

## MATERIALS AND METHODS

### Serum collection

This study was approved by the Institutional Ethics Review Committee of Foshan Fourth Hospital, Foshan, China. The written informed consent was obtained from each patient. COVID-19 patients were hospitalized and received routine treatment in Foshan Forth hospital during the period from 2020-1-25 to 2020-3-8 with variable stay time (**Table S1**). When the patients met the standard criteria of hospital discharge, the serum were collected, processed, and stored by standard protocol. As for the hospital discharge criteria, the vital points were normal body temperature, improved respiratory symptoms, lack of inflammation in the pulmonary imaging, and twice consecutively negative for nuclei acid testing. Sera of the control group from Lung cancer patients and healthy controls were collected from Ruijin Hospital, Shanghai, China. All sera were stored at −80°C until use.

### Antibody purification

S2 and N were biotinylated according to the manufacturer’s protocol. All the biotinylated proteins were incubated with Dynabeads^TM^ Myone^TM^ streptavidin T1 at room temperature for 1 hr, the excess proteins were removed by washing the magnetic beads with PBST (PBS containing 0.1% tween 20). The four biotinylated proteins, *i. e*., RBD, S1, S2 and N, were immobilized on the surface of magnetic beads, and labelled as RBD::magnetic beads, S1::magnetic beads, S2::magnetic beads and N::magnetic beads, respectively. To deactivate the potential live viruses in the sample, the serum from COVID-19 patients were incubated at 56 for 1 hr. As shown in **Fig. S1**, these sera were incubated with Dynabeads^TM^ Myone^TM^ streptavidin T1 to remove the antibodies that bind the free magnetic beads. The sera were incubated consecutively by the RBD::magnetic beads, S1::magnetic beads, S2::magnetic beads and N::magnetic beads at 4, each for 4 hrs. All the magnetic beads, containing the antibodies for each protein, were washed with PBST to remove the non-specific bindings. The antibodies on the beads were eluted with 50 mM glycine (pH2.8), and neutralized with Tris buffer (pH8.0). The quality and concentration of the protein enriched antibodies were checked and measured by SDS-PAGE. All the enriched antibodies were stored in −80 until in use. The same procedure was followed when enrich antibodies by biotinylated peptides.

### Identification of the binding motifs by AbMap

We followed the AbMap procedure as described previously (*20*) with slight modifications. Briefly, the 96-well PCR plates were blocked with PBST containing 3% BSA at 4 for 16 hrs. After washed with PBST, the plates were fulfilled with 200 µL PBST. Each well of the plates was loaded with 10 µL Ph.D.-12 phage display libraries. The samples, *i. e*., 1 µL for serum sample or α-His antibody and 400 ng for proteins enriched antibodies, were added per well. Blank control was set without the addition of sample, but Ph.D.-12 phage display library. The mixtures were incubated at 4 for16 hrs. Dynabesads^TM^ Protein G was added into each well to capture the antibody and phage complex. After incubated for 4 hrs at 4, the magnetic beads in each well were collected and washed. The beads in each well were suspended to 15 μL ultrapure water. After boiled at 98 for 10 min, the supernatant was collected.

To introduce the adapter sequence and unique barcode or index for each sample, two rounds of PCR were carried out on the phage lysate by Q5 hot start polymerase. The first round of PCR was performed by using primers XX-S5XX-23R and XX-N7XX-18 (5’-TCGTCGGCAGCGTCAGATGTGTATAAGAGACAGXXXXXXXX<u>GTGGTACCTTTCTATTCTCACTCT-3’</u>, and 5’-GTCTCGTGGGCTCGGAGATGTGTATAAGAGACAGXXXXXXXX<u>TTCAACAGTTTCGGCCGAACCT-3’</u>, where “XXXXXXXX” denotes a 8-nt barcode sequences, and the sequence with the underline was the specific primer for amplifying the corresponding nucleotide acid of the displayed peptide from the genome of phage, and the remaining sequence was the Illumina index). After checked by electrophoresis, all the PCR products were mixed. The PCR products were purified and used as the template for the second round PCR. In the second round PCR, the unique indexes of Illumina next generation sequencing (NGS) were introduced for each mixture. After purified by gel extraction, the final sample was ready for NGS.

According to the index and barcode combination, the results of NGS were split and assigned to each sample. For each sample, the data of NGS were trimmed further, and only sequence of 36 bp was left, which corresponded to the 12-mer displayed peptide. All the sequences were translated into peptides, and the frequency of each peptide was counted. If stop codon appeared in the translation process, the corresponding sequence was removed. The enrichment factor of each peptide from the sample were calculated. The reverse enrichment factor of each peptide was also calculated, of which the maximum value was set as the cutoff of the corresponding sample. The peptides, with the enrichment factors lager than the cutoff was determined as the hits. These hits were subjected for MEME analysis, significant motifs were called by setting E value less than 0.05 as the cutoff (*40*). The significant motifs were matched to SARS-CoV-2 protein/s by FIMO used default settings (*36*).

### Map the epitopes to the 3D structures of the proteins

A Spike protein structure (PDB ID: 6X6P) was used to analyze the location of the epitopes on the 3D structure. RBD/ACE2 complex structure (PDB ID: 6MOJ), RBD/CR3022 complex (PDB ID: 6W41), RBD/S309 complex (PDB ID: 6WPT), RBD/P2B-2F6 complex (PDB ID: 7BWJ), RBD/BD23 complex (PDB ID: 7BYR) and RBD/CB6 complex (PDB ID: 7C01) were used to analyze the location of the epitopes on these protein complexes. All the structural analyses were processed by using Pymol.

### Validation by peptide microarray

Peptides were synthesized and conjugated to BSA by GL Biochem Ltd. (Shanghai, China). The concentration of each peptide was diluted to 0.5 mg/mL, 0.25 mg/mL and 0.125 mg/mL with PBS containing 40% glycerin, respectively. According to the pattern showed in **Fig. S6**, all the peptides, with extra controls, such as the blank control, human IgG, human IgM and land marker Cy5-labelled donkey anti-human IgM, were printed on the surface of PATH slide with 2 × 7 identical subarrays by using Super Marathon printer (Arrayjet, UK). The peptide microarray was stored in −80°C until use.

The assay on the peptide microarray was similar to our previous study (*15*). Briefly, the peptide microarrays were blocked with PBS containing 3% BSA. The slides were washed with PBST, and assembled with a 14-chamber rubber gasket. Serum and the enriched antibodies were diluted by 1:200 in incubation buffer and incubated with the array for 2 hrs at room temperature. After three washes with PBST buffer, the arrays were incubated by Cy3-conjugated goat anti-human IgG and Cy5-conjugated donkey anti-human IgM for 1 h at room temperature at 1: 1000 dilution in 1x PBST. The microarrays were washed three times with 1×PBST, dried by centrifugation and scanned by LuxScan 10K-A (CapitalBio Corporation, Beijing, China). The fluorescent intensity was extracted by GenePix Pro 6.0 software (Molecular Devices, CA, USA).

### Statistical Analysis

The fluorescent intensity was extracted by GenePix Pro 6.0 software (Molecular Devices, CA, USA). The signal intensity was defined as the mean of the foreground of each spot, and then averaged the three spots of each peptide. All the Signal intensities were analyzed by the software GraphPad Prism version 7.0.

## SUPPLEMENTAL FIGURE LEGENDS

**Figure S1.**
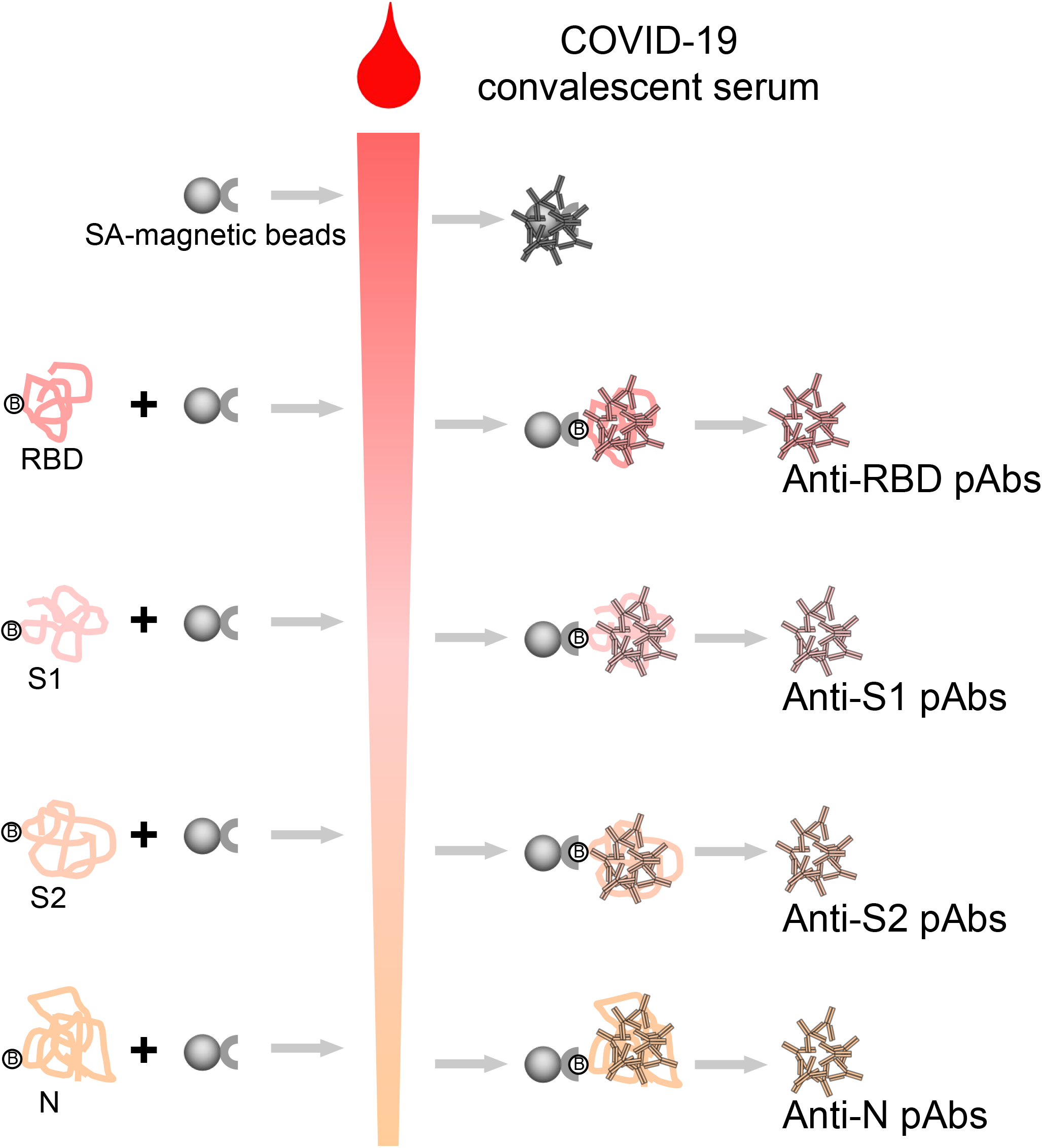
Protein specific antibodies enrichment from COVID 19 convalescent sera. COVID-19 convalescent sera were used as input. Terminal biotinylated proteins were used as baits to enrich specific antibodies. SA conjugated magnetic beads were adopted to isolate the antibodies.

**Figure S2.**
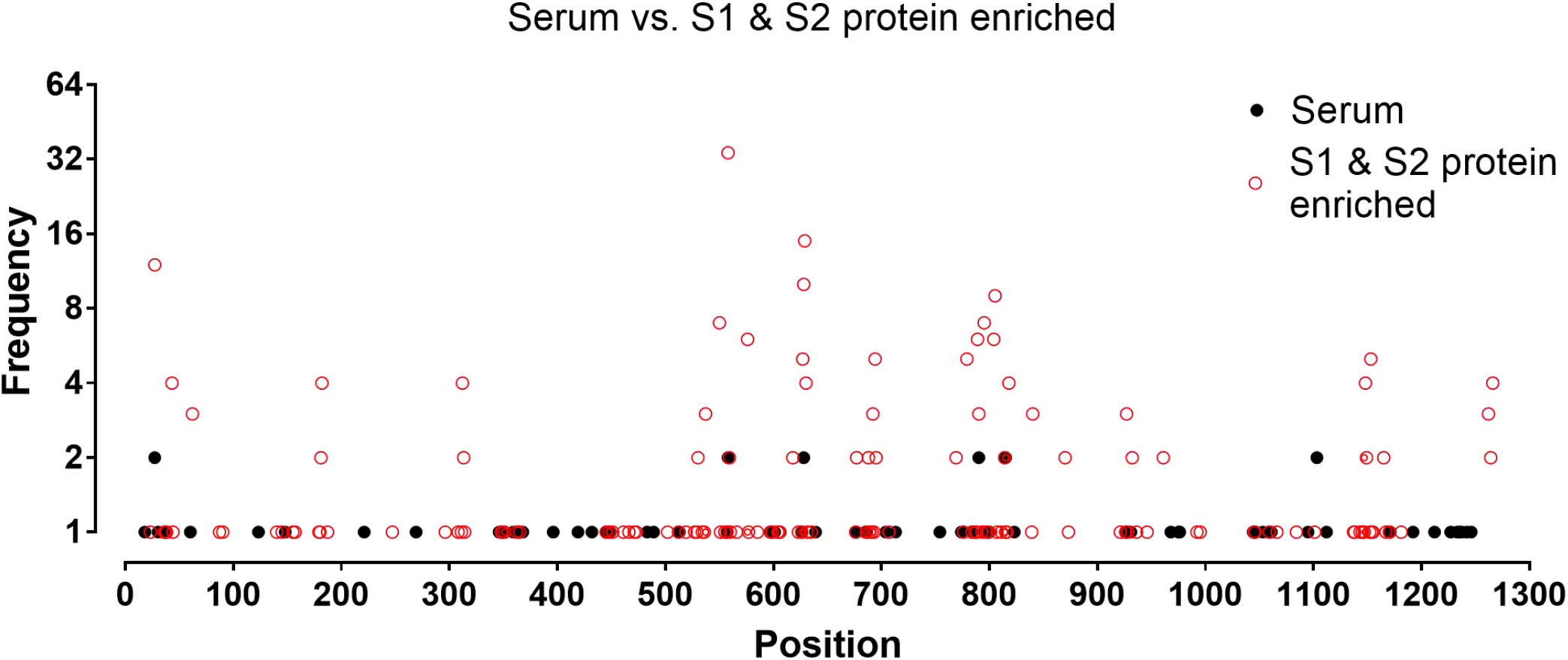
Protein based antibody enrichment is necessary for AbMap. S protein specific epitopes identified from sera directly (black dot), and S1+S2 protein enriched antibodies (red dot).

**Figure S3.**
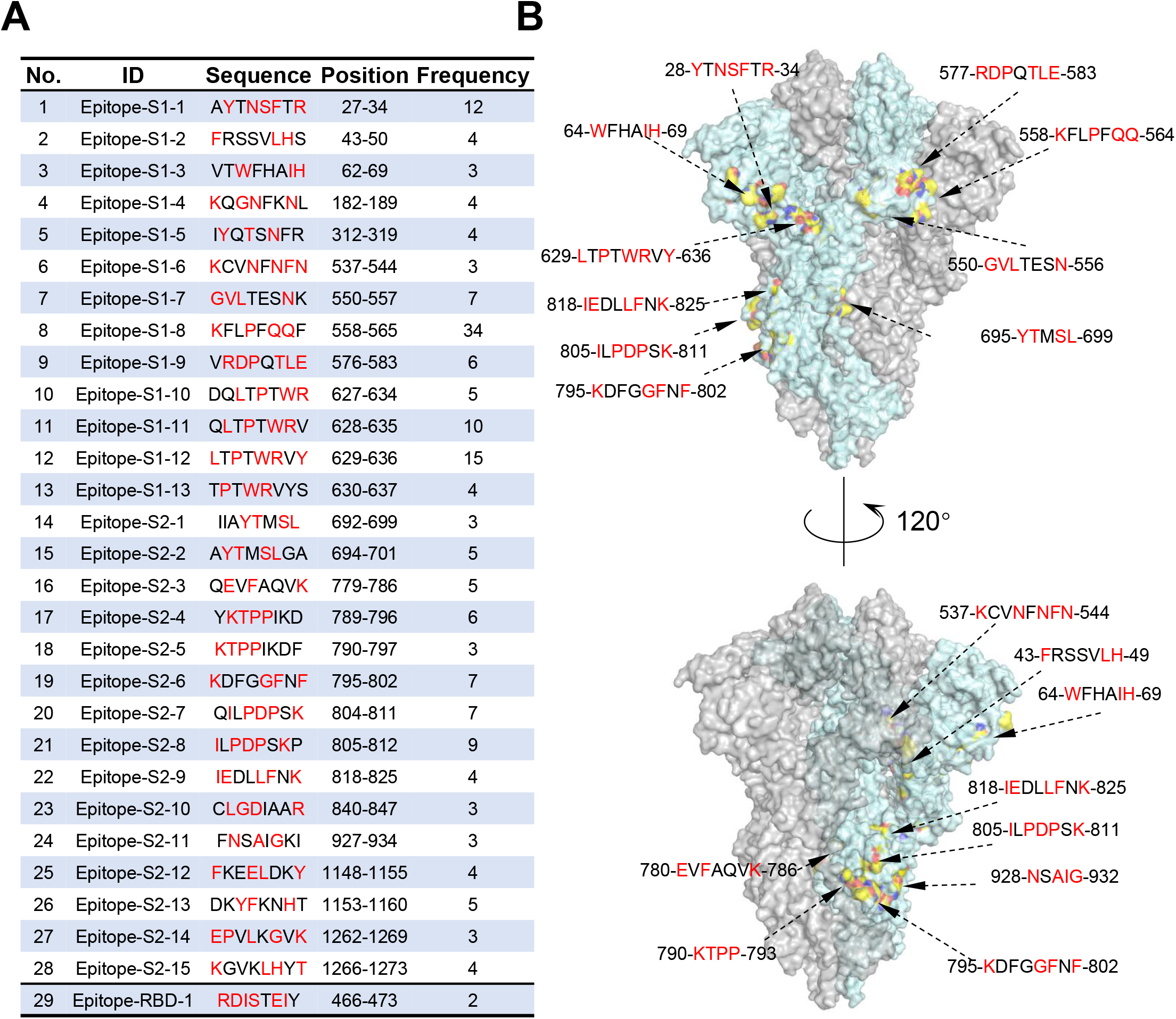
Detailed information of the significant epitopes on S protein. (A) The list of the significant epitopes (frequency > = 3), (B) The distribution of the significant epitopes (frequency> = 3) on the trimer 3D structure of the S protein. Red marked amino acids represent critical residues of epitopes.

**Figure S4.**
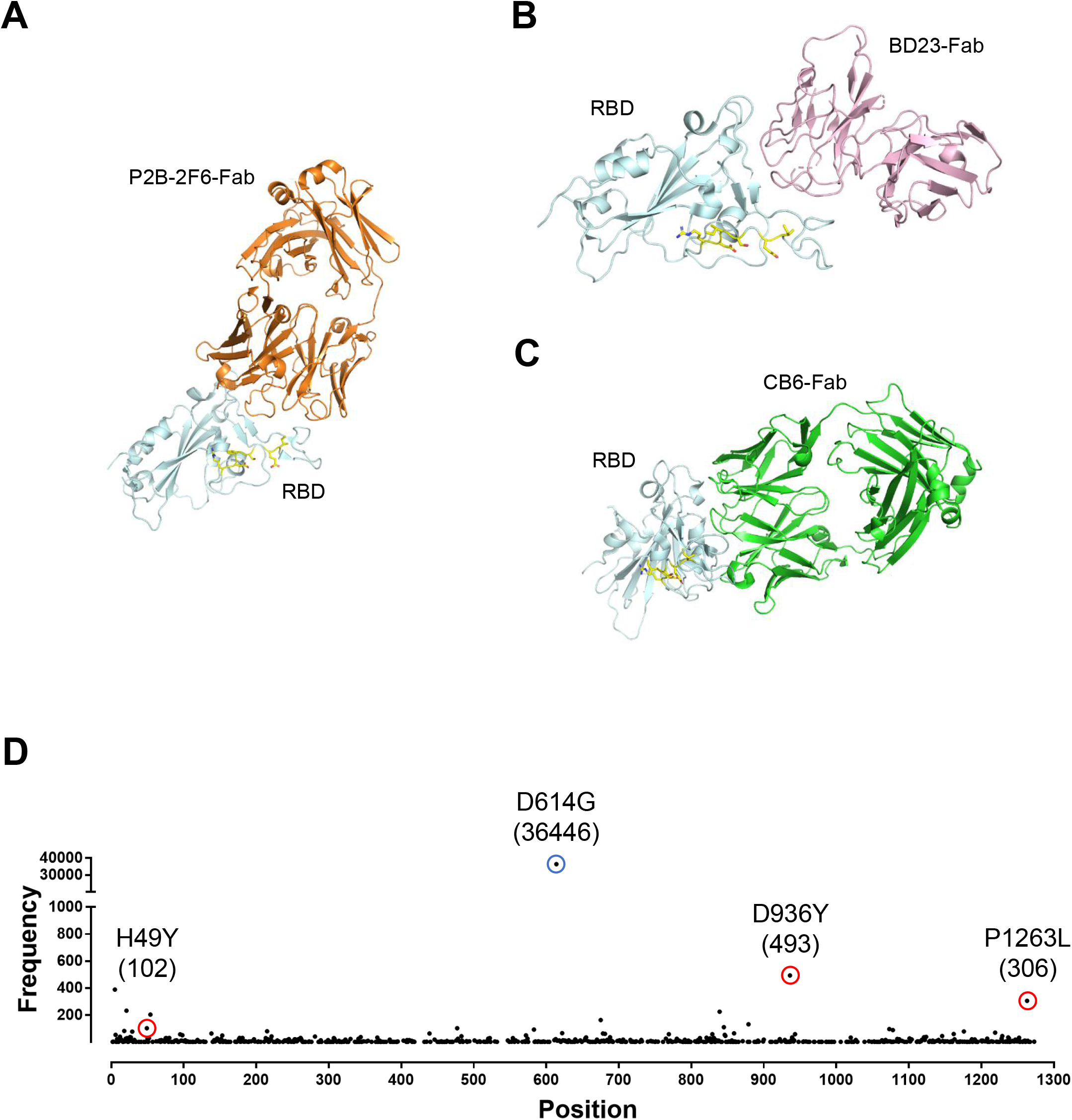
Additional structure analysis of a significant epitope on RBD and Naturelly existing mutants of Spike protein. (A-C) The key residues locate adjacent to but not at the binding interfaces of RBD by other three highly potent neutralization antibodies, P2B-2F6-Fab (A), BD23-Fab (B) and CB6-Fab (C). (D) The Substitution mutants of Spike protein collected by China National Center for Bioinformation (CNCB) were plotted alongside the Spike protein with the mutated frequency. (The data were collected by 2020–8–26)

**Figure S5.**
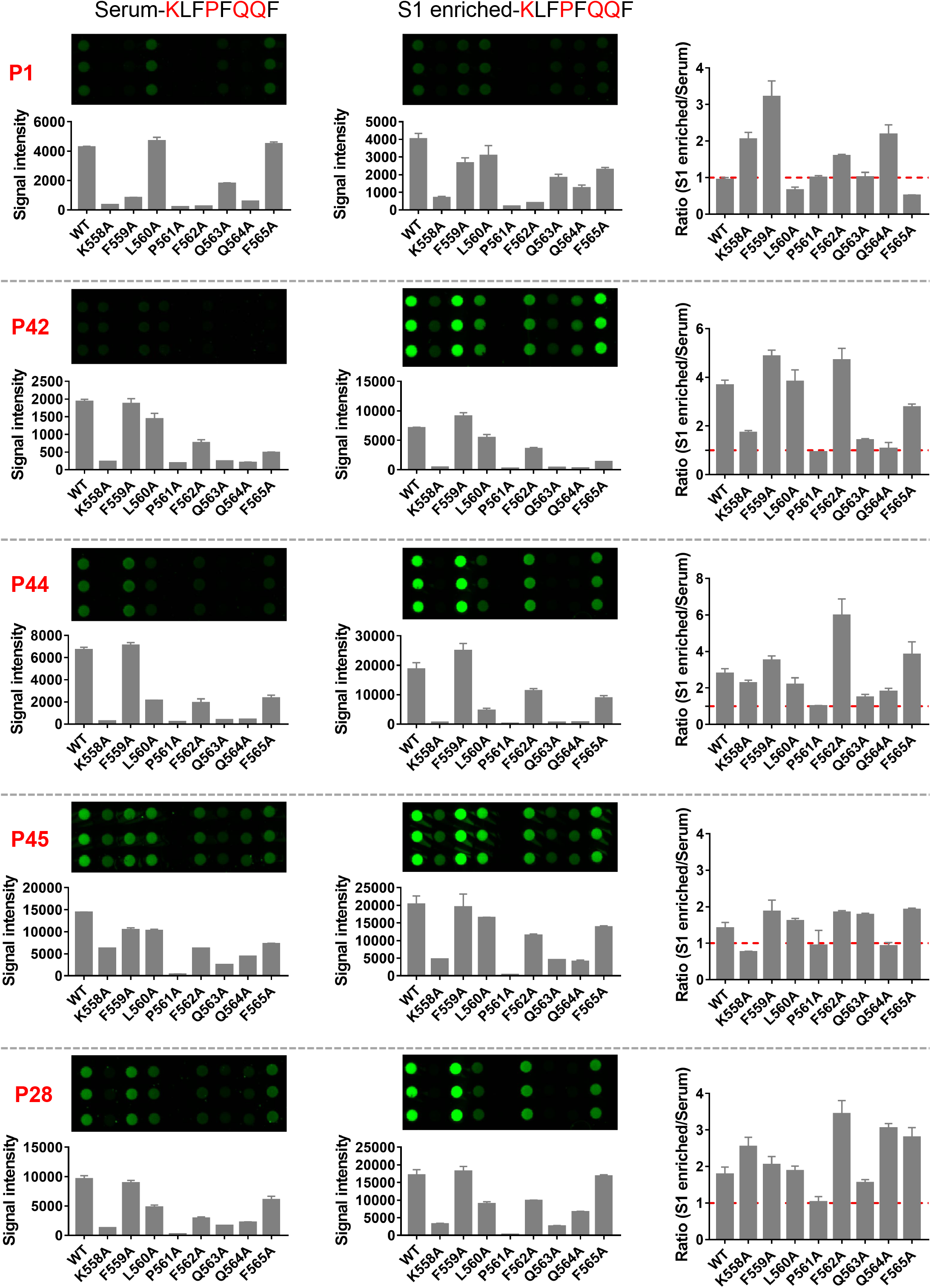
Validation of epitopes by a peptide microarray: the comparison of sera and protein enriched antibodies. Peptide 1 (Pep 1), which corresponds to Epitope-S1–8 (KLFPFQQF) was selected as an example. Sample with red label indicates the corresponding epitope was identified from this sample by AbMap. The ratio of signal_intensity (S1 enriched)/ signal_intensity (serum) for each residue was plotted (right).

**Figure S6.**
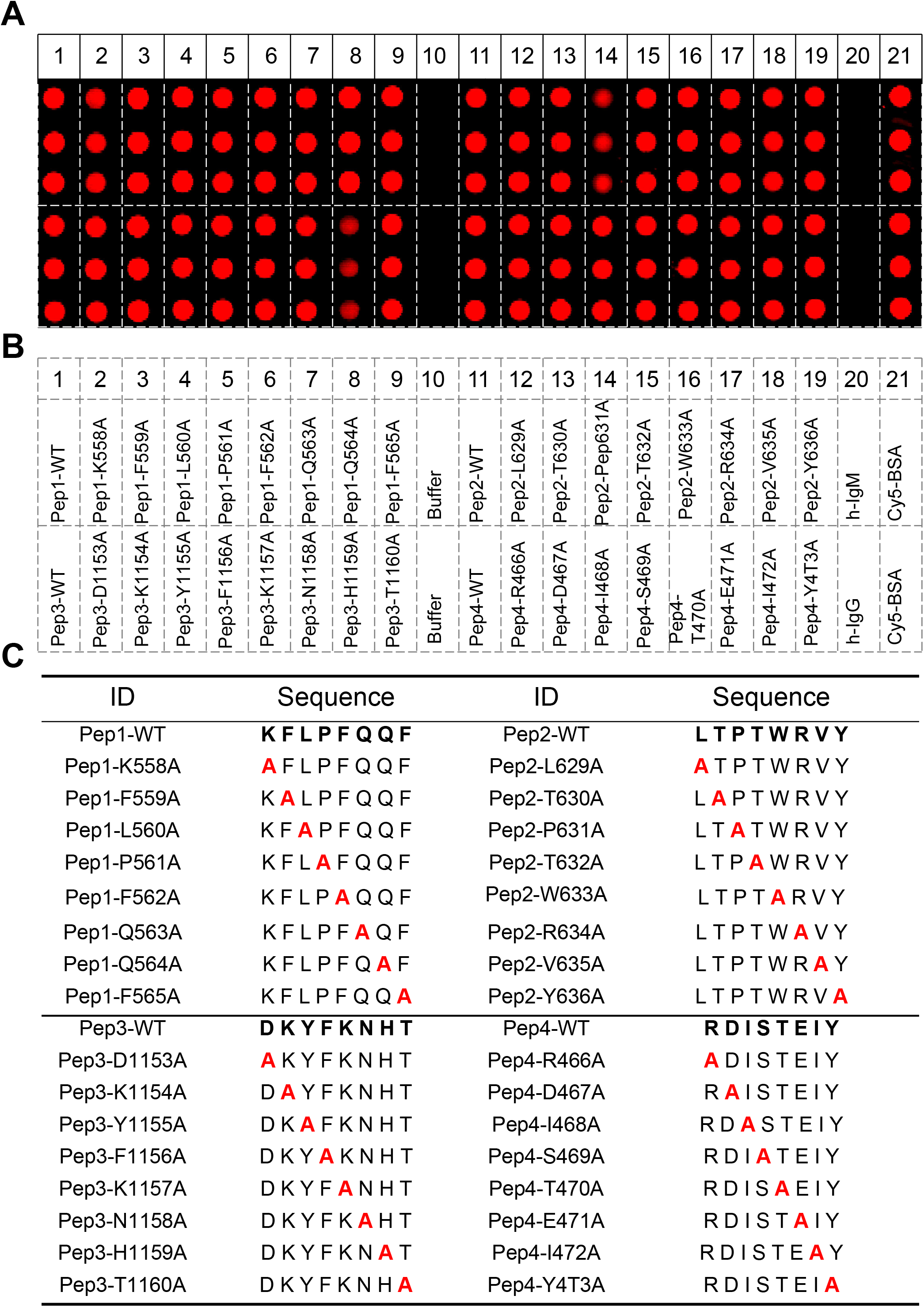
The layout of the peptide microarray of 4 significant epitopes. (A) Quality control of the peptide microarray. All the peptides were chemically synthesized and conjugated to BSA. The BSA conjugated peptides were printed and immobilized on the microarray. The microarray was stained with an anti-BSA antibody and followed with a Cy5 conjugated secondary antibody. (B) The layout of the peptide microarray. (C) The sequences of the peptides that were included on the peptide microarray.

**Table S1.**
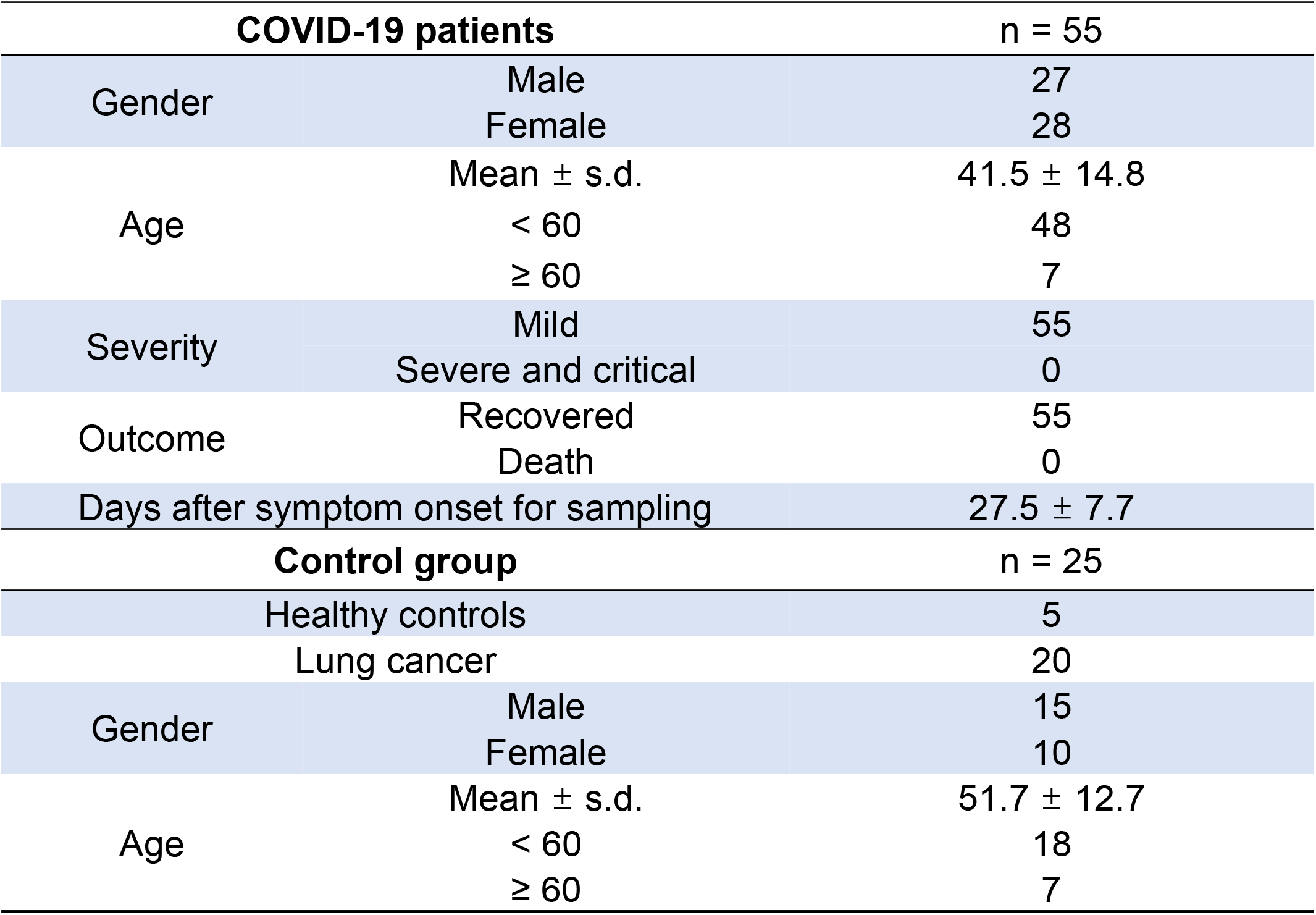
Serum samples tested in this study.

| **COVID-19 patients** |  | **n = 55** |
| --- | --- | --- |
| Gender | Male | 27 |
|  | Female | 28 |
| Age | Mean ± s.d. | 41.5 ± 14.8 |
|  | < 60 | 48 |
| | ≥ 60 | 7 |
| Severity | Mild | 55 |
|  | Severe and critical | 0 |
| Outcome | Recovered | 55 |
|  | Death | 0 |
| Days after symptom onset for sampling | | 27.5 ± 7.7 |
| **Control group** |  | **n = 25** |
| Healthy controls |  | 5 |
| Lung cancer |  | 20 |
| Gender | Male | 15 |
|  | Female | 10 |
| Age | Mean ± s.d. | 51.7 ± 12.7 |
|  | < 60 | 18 |
| | ≥ 60 | 7 |

**Table S2.**
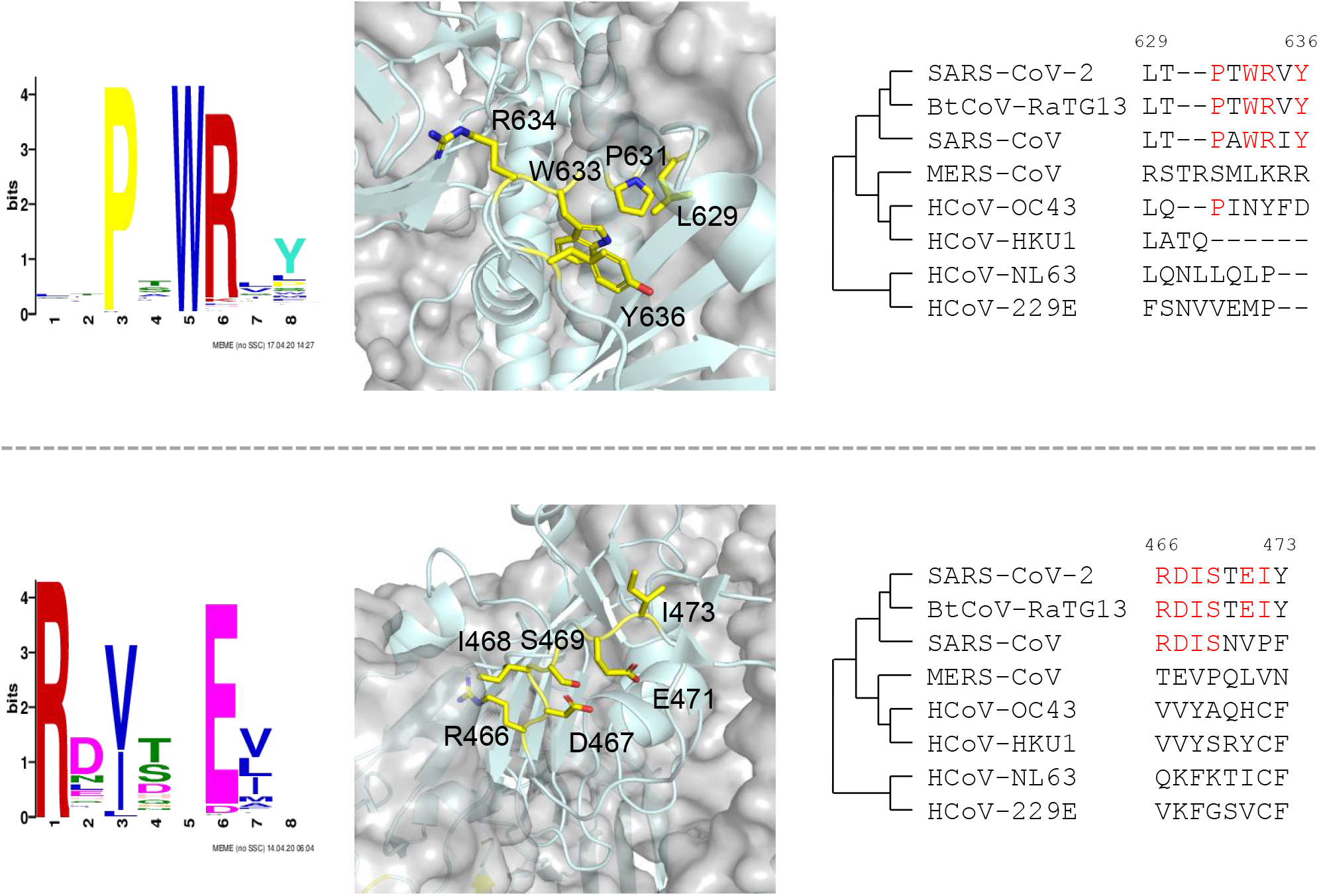
All the identified motifs.

**Table S3.**
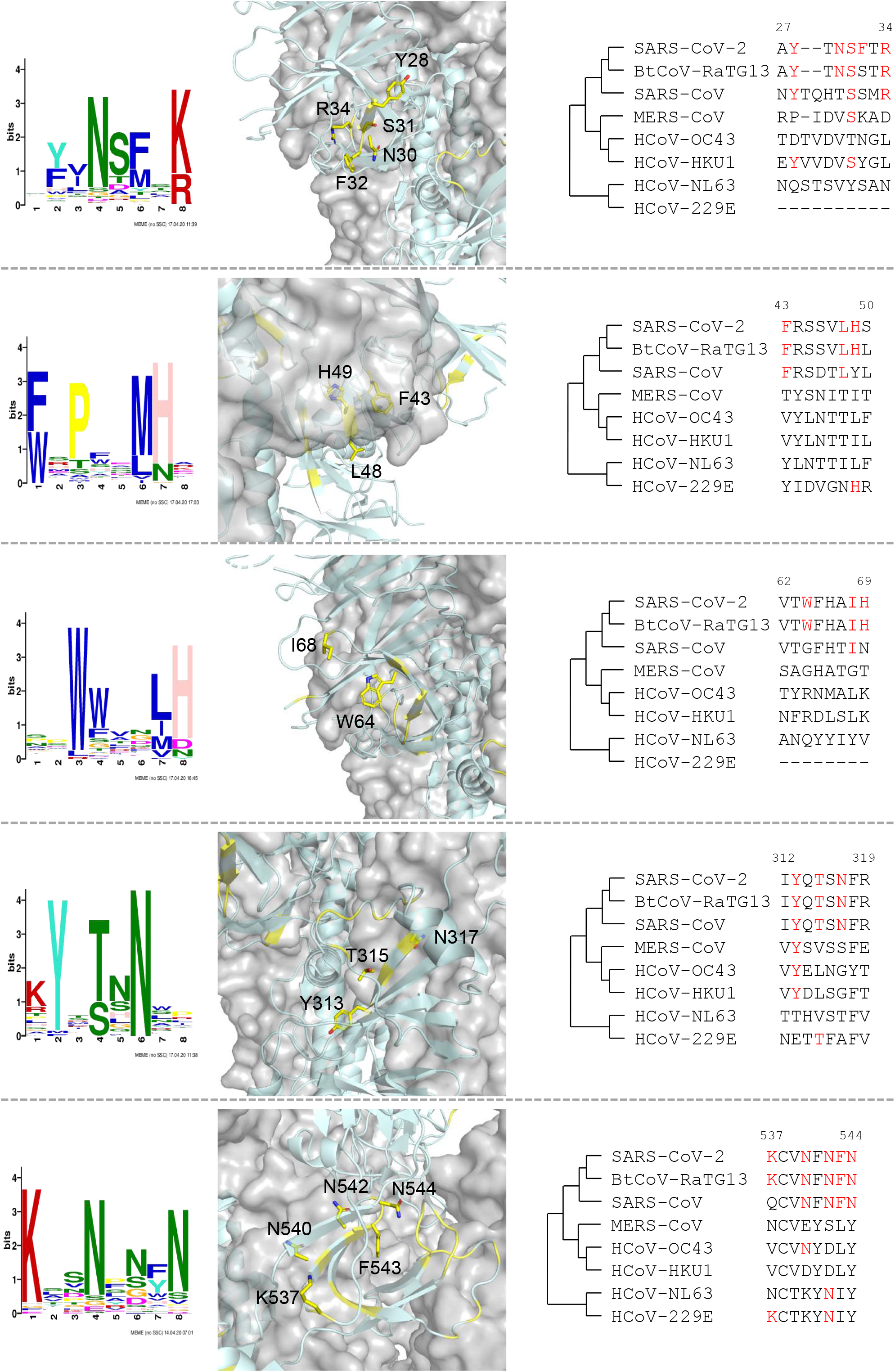
The identified epitopes that match the sequences of SARS-CoV-2 proteins.

**Table S4.**
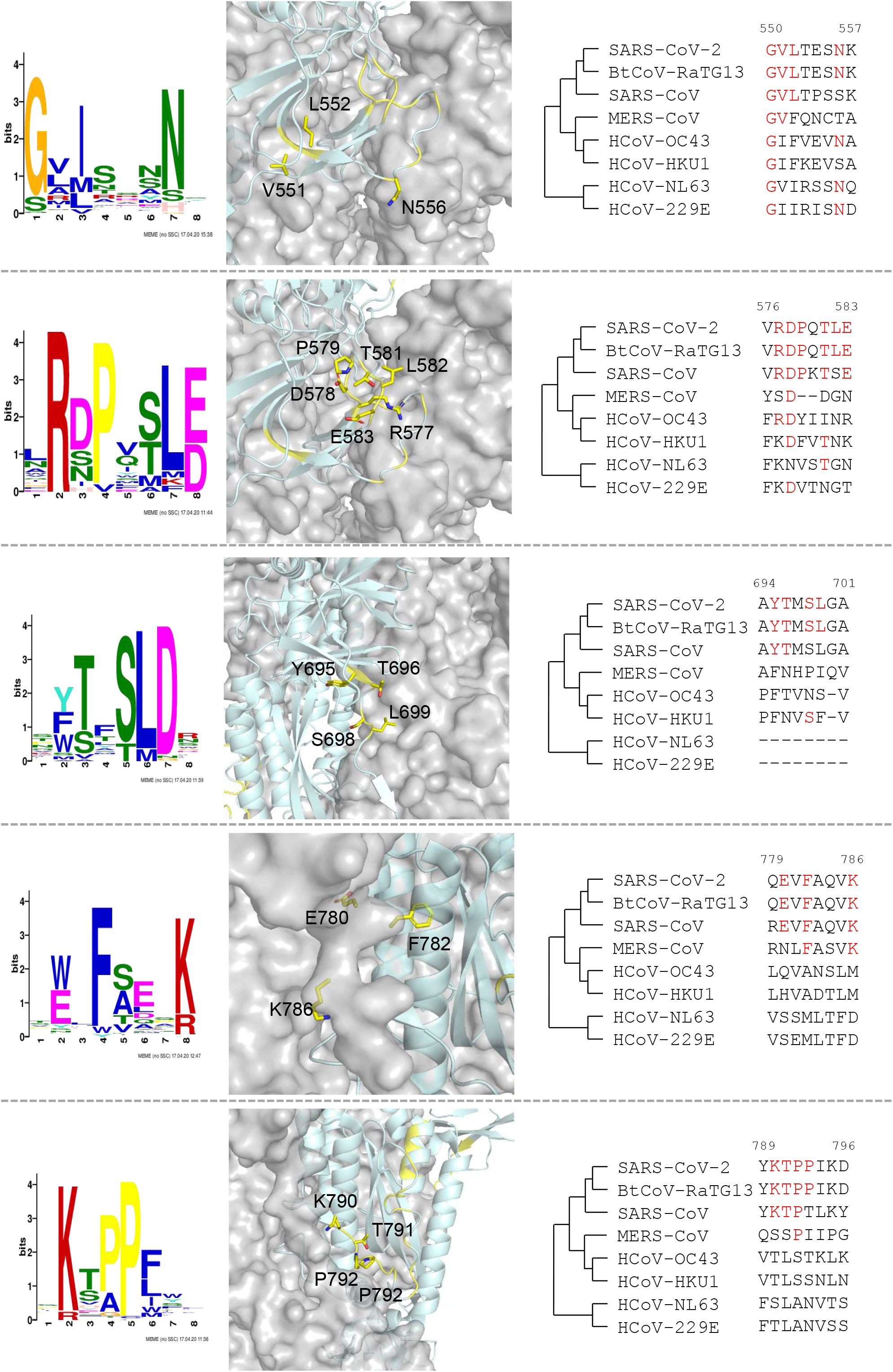
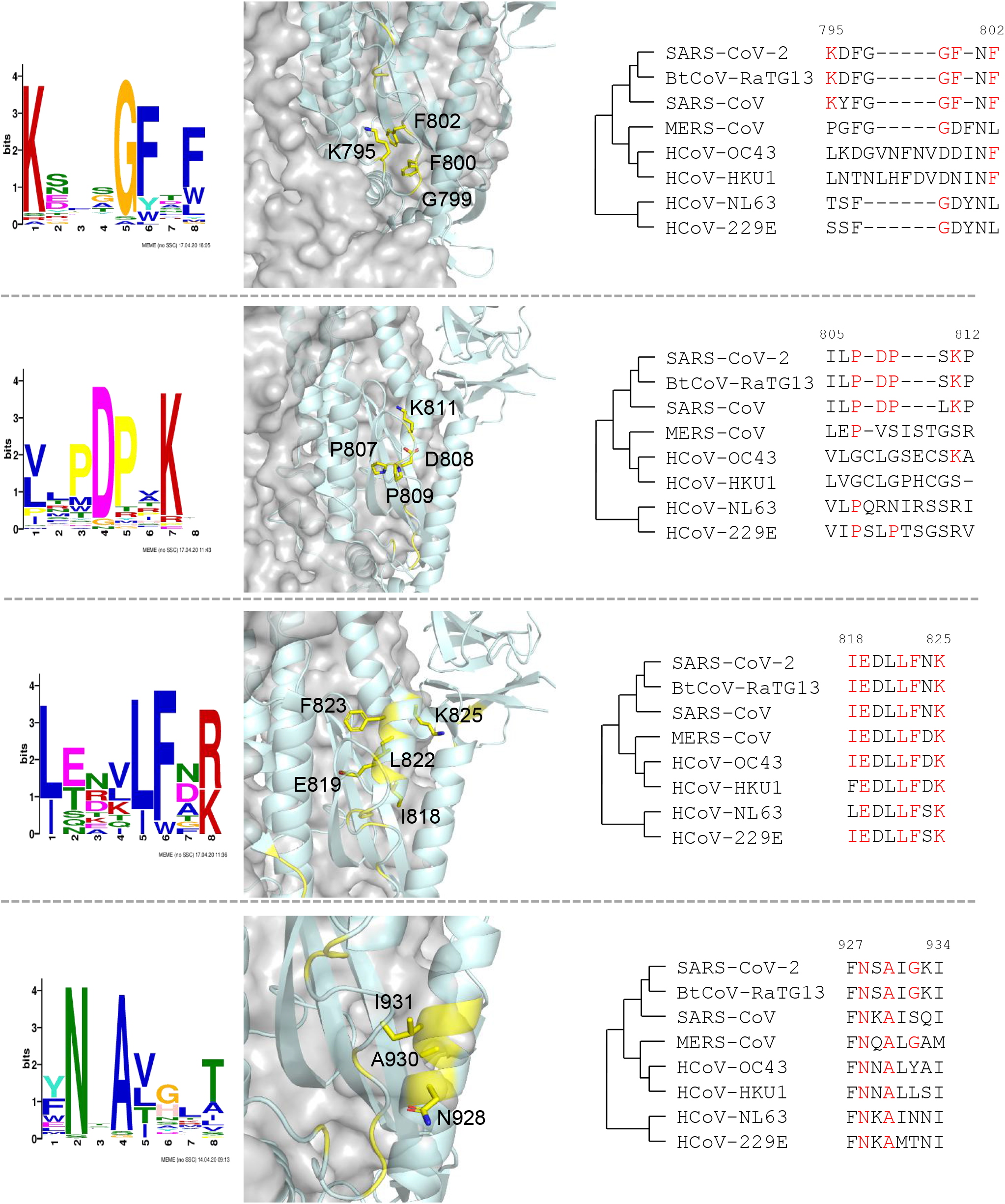
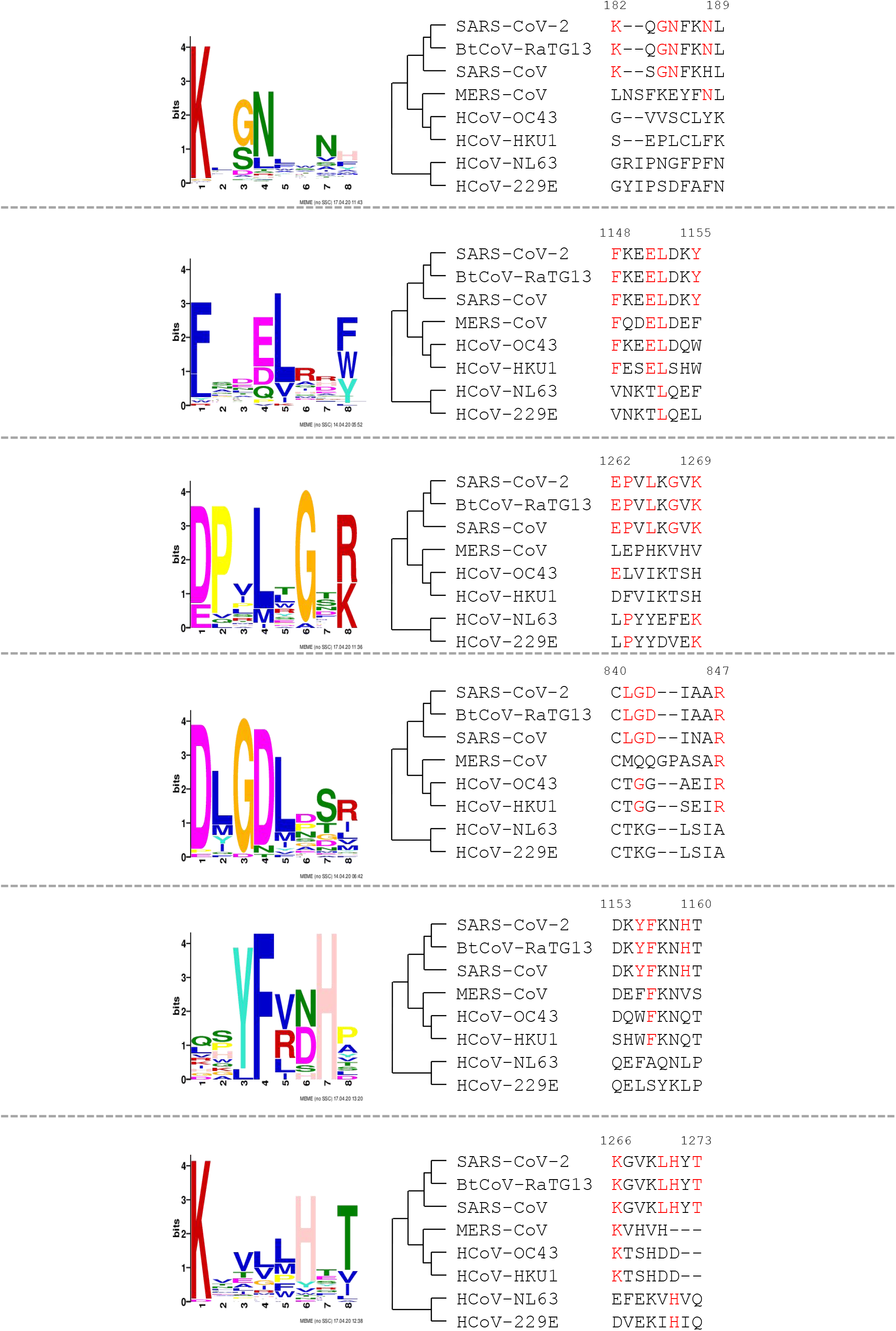
Structure and homology analysis of significant epitopes. Two significant epitopes. The epitopes were matched to S protein, critical epitope residues were labeled as yellow. Homology analysis of the epitope among three deadly coronaviruses, four common human coronaviruses, and bat coronavirus BtCoV-RaTG13.

